# COVID19 vaccines as boosters or first doses: Simulating scenarios to minimize infections and deaths

**DOI:** 10.1101/2024.04.12.24305705

**Authors:** Omar El Deeb, Joseph El Khoury Edde

## Abstract

Public health authorities face the issue of optimal vaccine distribution during spread of pandemics. In this paper, we study the optimal way to distribute a finite stock of COVID-19 doses between first or second doses for unvaccinated individuals and third doses (booster shots) for fully vaccinated individuals. We introduce a novel compartmental model that accommodates for vaccinated populations. This Booster model is implemented to simulate two prototypes of populations: one with a highly infected and highly vaccinated proportion, and another with a low infected and vaccinated percentage. We namely use sample data from Russia and Djibouti respectively.

Our findings show that, to minimize the deaths for the first type of populations, around one quarter of the vaccines should be employed as booster shots and the rest as first and second doses. On the other hand, the second type of populations can minimize their number of deaths by mainly focusing on administering the initial two doses, rather than giving any booster shots. The novel Booster model allows us to study the effect of the third dose on a community and provides a useful tool to draw public policies on the distribution of vaccines during pandemics.

## 1. Introduction

Throughout history, mankind has faced numerous pandemics, causing health, economic, and political crises around the globe. Although these pandemics resulted in hundreds of millions of deaths, in hindsight, these catastrophes motivated research that gave rise to major breakthroughs in the related public health and medical fields. The Coronavirus (COVID-19) was proclaimed a global pandemic in March of 2020 after spreading worldwide, starting from China where the first case appeared in December 2019, then Italy and Iran, and subsequently the whole world. The most common symptoms of infection included fever or chills, cough, shortness of breath or difficulty breathing, fatigue, muscle or body aches, headache, loss of taste or smell, and so on. This is caused by the airborne SARS-CoV-2 virus, which is responsible for over 692 million confirmed cases and 6.9 million deaths globally as of July 2023 [1]. Governments around the world were obliged to enforce strict measures to mitigate the spread of this contagion, resulting in adverse effects on people’s lives and routines. Since the beginning of the COVID-19 pandemic, the SARS-CoV-2 virus has mutated numerous times. In most cases, the changes made to the genetic material led to little or no impact on the properties of the virus itself, such as its severity and transmission rate. However, sometimes these mutations can lead to the formation of variants of concern (VOCs). VOCs are classified as mutations of viruses that have proven to increase the transmissibility of the virus, increase the severity of the disease, and decrease the effectiveness of vaccines and other medical treatments [2]. There have been multiple cases of VOCs originating from different countries around the world at different times, such as the Alpha variant from the United Kingdom in September 2020, the Delta variant from India in October 2020, and the Omicron variant from South Africa in November 2021 [3].

The pharmaceutical industry has developed multiple forms of vaccines approved by the World Health Organization along with other national health agencies. Each vaccine uses a specific approach to enhance the immune system’s response against the virus. There are genetic vaccines that contain a segment of the virus, inactivated vaccines that contain a killed sample of the Sars-CoV-2 virus, attenuated vaccines that contain a weakened virus, and protein vaccines that contain different protein fragments of the virus [4]. Further research is currently investigating different forms of vaccines, such as dry vaccines that can be inhaled, and antiviral drugs that enhance the body’s ability to fight the Sars-CoV-2 virus [5]. Clinical studies showed that Anti-body therapy is effective for severe cases of COVID-19, as there was a 20% higher chance of survival among patients. However, this method is considered highly costly and remains in short supply [6]. Furthermore, previous studies have shown that the effectiveness of the Moderna vaccine is around 93% two weeks after the second dose; similarly, 4 months after full vaccination, the effectiveness remains around 92%. While the Pfizer-BioNTech vaccine starts off with 91% efficacy 2 weeks after the second dose, it slightly loses effectiveness with time to reach 77% efficacy 4 months after the second dose. Finally, Johnson & Johnson, the one-dose vaccine, was shown to have a 71% vaccine effectiveness two weeks after vaccination, and a 68% effectiveness one month after vaccination [7].

Attempting to predict the number of active cases at any point in the future for any given set of initial conditions is important since a recent study has shown that approximately 2% of the COVID-19 cases were admitted to the hospital [8]. Therefore, if the number of cases increases drastically, then the number of infected individuals in need of hospitalization due to COVID- 19 may exceed the hospital’s capacity to admit these patients.

The aim of this study is to predict the spread of infectious diseases, specifically COVID-19, and explore the optimal use of a limited number of vaccines by examining various proportions of vaccine doses that can be used for different initial conditions. The study investigates the expected number of active cases, deaths, as well as the number of hospitalized patients due to the COVID-19 pandemic during which the available vaccines are administered as boosters for previously vaccinated individuals or as first and second doses for unvaccinated individuals. The results of this study are important as the world still faces the imminent threat of the emergence of new variants and new viruses that may cause a resurgence in the pandemic, with a limited supplyof vaccines that would ultimately be available to fight them. Prioritizing the administration ofvaccines for a specific group or another would be a very important factor to draw publicpolicies by health authorities across the globe.

## 2. Literature Review

Mathematical modeling of infectious diseases is a lively interdisciplinary field of research that brings together researchers from Biology, Epidemiology, Mathematics, Statistics, Physics, and Medicine. These models allow researchers to forecast, predict, and quantify the uncertainty of their forecasts.

The first compartmental model used was the SIR model, created by Kermack and McKendrick in 1929, which splits the population into 3 compartments: Susceptible (S), Infected (I), and Removed (R) [9]. This model was later modified to consider other factors such as undetected infections, environmental factors, traveling, lockdown, non-pharmaceutical treatments, and vaccinations. Environmental factors are incorporated into the model to understand their impact on the viability and transmission of pathogens, allowing researchers to simulate seasonal variations in disease spread. To account for travel effects, SIR models are extended to spatial models, dividing populations into distinct geographical regions, and considering patterns of human movement between these areas. This integration of travel data enables the assessment of how population mobility influences the spread of infectious diseases. Moreover, SIR models are invaluable for evaluating the impact of lockdown measures on disease transmission. By modeling interventions like social distancing, quarantines, and the closure of public spaces, researchers can assess the effectiveness of various strategies in slowing down the spread of the disease. Adjusting parameters, such as contact rates between individuals, based on the severity and duration of lockdown measures, allows for the simulation of different scenarios and predictions about their influence on the epidemic’s course [10–16]. Additionally, SIR models are employed to analyze the effects of vaccination on disease dynamics. Researchers can simulate the vaccination of susceptible individuals, altering the parameters to reflect the coverage, efficacy, and timing of vaccination campaigns. This helps assess the potential impact of vaccination in reducing the overall transmission and severity of the disease within the population [17–24].

While studying the durability of the vaccine, it was found that those who were successfully immunized after vaccination had a strong vaccination for 6 months, or 180 days, after vaccination [25]. For that reason, countries such as the United States and France have introduced a third shot, or a booster shot, to reimmunize those who have lost their immunity [26–28]. Recent studies revealed a dose-dependent reduction in the percentage of infected individuals within the vaccinated population. The simulation results exhibited a close alignment with real-world data on infected patients, affirming the appropriateness of the model [29].

## 3. Methods: The *Booster* Model

One of the major topics that needs to be studied in detail is the COVID-19 third dose or booster shot. As mentioned previously, the immunity gained through vaccination is not permanent. In this model, it will be taken as 6 months, or 180 days in accordance with the latest studies.

However, this immunity can be reacquired through the administration of a single booster shot. Our study introduces a novel approach with the objective of minimizing deaths in two distinct population types. So, a new question arises: Which vaccination scheme leads to less deaths: giving unvaccinated individuals 2 doses, or giving double the number of people, a booster shot? This issue could also be similarly addressed on the global level, given the huge disparities in availability of vaccines among different countries. For an ideal scenario of global vaccine equity, would it be more efficient to continue supplying booster shots in countries that already achieved high percentages of vaccination, or to prioritize allocating these resources to spread first doses among populations that didn’t have enough access into them yet?

To answer this question, the *Booster* model was introduced. It is a compartmental modelmade up of 7 compartments which are: Susceptible (S), Susceptible vaccinated (SV), Susceptible with lost immunity (SL), Infected (I), Infected vaccinated (IV), Recovered (R), and Dead (D). The *Booster* model can be represented with the following system of ordinary differential equations:

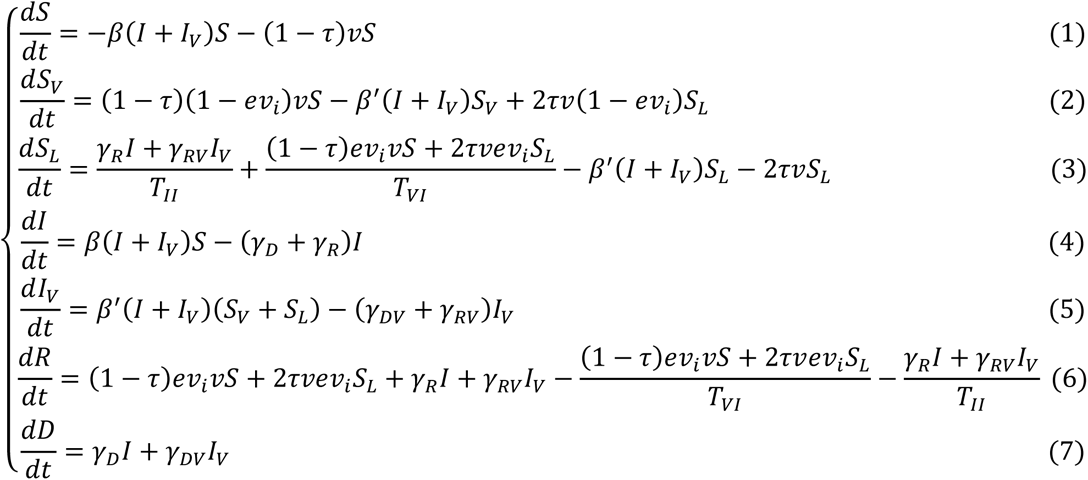

Where S is the fraction of the population, which is unvaccinated and susceptible to the virus, SV is the vaccinated susceptible population, I is the unvaccinated infected population, IV is the vaccinated infected population, R is the recovered population, and D is the dead population.

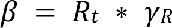 is the effective contact rate between infected individuals (I+IV) and the unvaccinated susceptible individuals S; likewise, 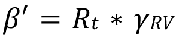 is the contact rate between infected individuals and the vaccinated susceptible individuals SV. *e* is the efficiency of the vaccine, 𝑣_𝑖_ is the maximum possible vaccine intake and *v* is the abundancy or roll out rate of the vaccine.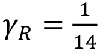 and 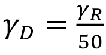 are the average rate of recovery and death of unvaccinated individuals respectively.

Similarly, 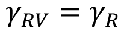 and 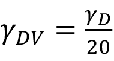 are average rates of the recovery and death of vaccinated individuals respectively. 𝑇𝐼𝐼 = 360 𝑑𝑎𝑦𝑠 and 𝑇𝐼𝑉 = 180 𝑑𝑎𝑦𝑠 are the average lengths of immunity after infection and vaccination respectively. Finally, 𝜏 is the proportion of vaccine doses used as booster shots. For example, if 𝜏 = 0.75, then three quarters of the vaccines are booster shots,while the remaining 25% are used as first or second doses.

Notice in the 3^rd^ and 6^th^ equation, there is 2𝜏𝑣𝑆𝐿 rather than 𝜏𝑣𝑆𝐿. This is because to give the booster shot to people who lost their immunity, only one does is required. Contrarily, to fully vaccinate unvaccinated individuals, 2 doses are need. As a result, half the doses are required to give booster shots, therefore, the rate at which booster shots are given to the public should be twice the rollout rate to the unvaccinated population, thereby explaining the presence of a factor of 2.

To visualize the interaction between different compartments, we graphically illustrate them in Fig. (1).

In order to apply the model to real life scenarios, two populations will be taken as prototypes, Russia and Djibouti. Russia will be used to simulate the dynamics of COVID-19 in a population with a relatively high infection and vaccination rate, while Djibouti will be used to model COVID-19 spread in a population with low infection and vaccination rate.

**Figure 1.**
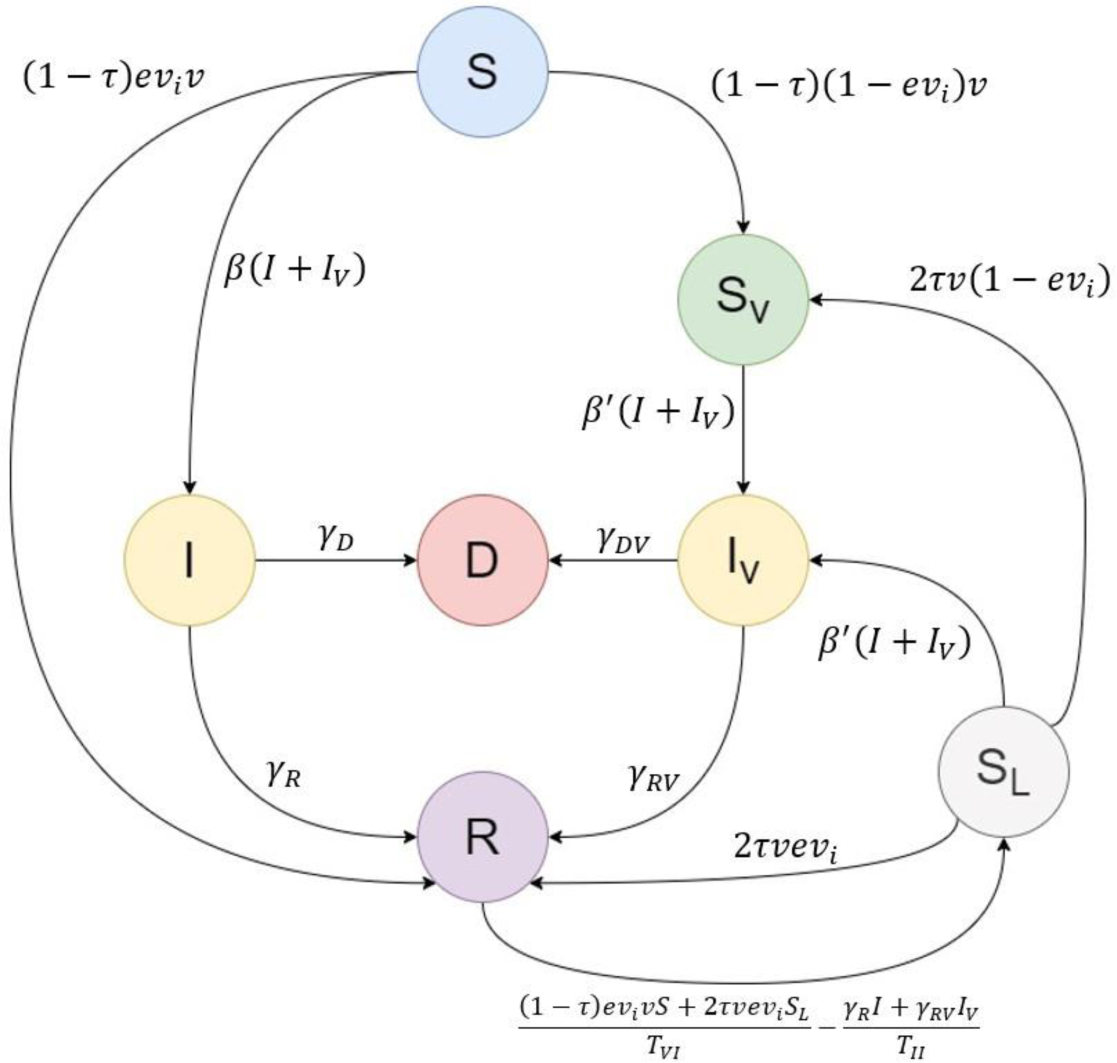
Schematic diagram showing the 7 compartments of the Booster model: Susceptible (S), Susceptible Vaccinated (SV), Susceptible with Lost immunity (SL), Infected (I), Infected Vaccinated (IV), Recovered (R), Dead (D) - along with the transfer dynamics between them.

Throughout the study, nine vaccination schemes were tested with vaccine efficacies (*e*) ranging between 55% and 92%, and vaccine abundancy or rollout rate (*v*) ranging between 0.1% and 1.5%. The schemes tested were: *e*=92% and *v*=0.1% - *e*=92% and *v*=0.3% - *e*=92% and *v*=0.5% - *e*=72% and *v*=0.3% - *e*=72% and *v*=0.5% - *e*=72% and *v*=0.7% - *e*=55% and *v*=0.7% - *e*=55% and *v*=1% - *e*=55% and *v*=1.5%, with vaccine intake assumed to be 𝑣_𝑖_ = 90% for all scenarios.

## 4. Results and Discussions

In mid-February 2022, there was a total of 2,668,036 active cases in Russia. Therefore, with a total population of 143.4 million, the percentage of the infected individuals was 1.84%. At that time, COVID-19 was the cause of 340,248 deaths in Russia, or 0.23% of the Russian population. In mid-February, there were 71.32 million individuals, or 49.74% of the Russian population, who were fully vaccinated, but 6 months prior, there were 32.43 million (22.62%) double vaccinated individuals; therefore 22.62% of the Russian population have probably lost their immunity after vaccination, leaving 27.12% that are currently immune [30, 31].

According to the data obtained in Fig. (2), if the vaccination scheme consists of vaccines with a high efficacy (*e* = 92%), it is better to use all the vaccine doses to fully vaccinate unvaccinated individuals. This can be seen in the top row of the figure above, as the curve resulting with the lowest number of cumulative deaths was achieved with a value of 𝜏 = 0. However, when the efficacy of the vaccine decreases, the significance of the third dose increases. Hence, if a vaccine has a low efficacy (*e* = 55%), the optimal value of tau is 𝜏 = 0.25, and if a vaccine has a moderate efficacy (*e* = 72%), both 𝜏 = 0 and 𝜏 = 0.25 were equally effective in decreasing mortality.

**Figure 2.**
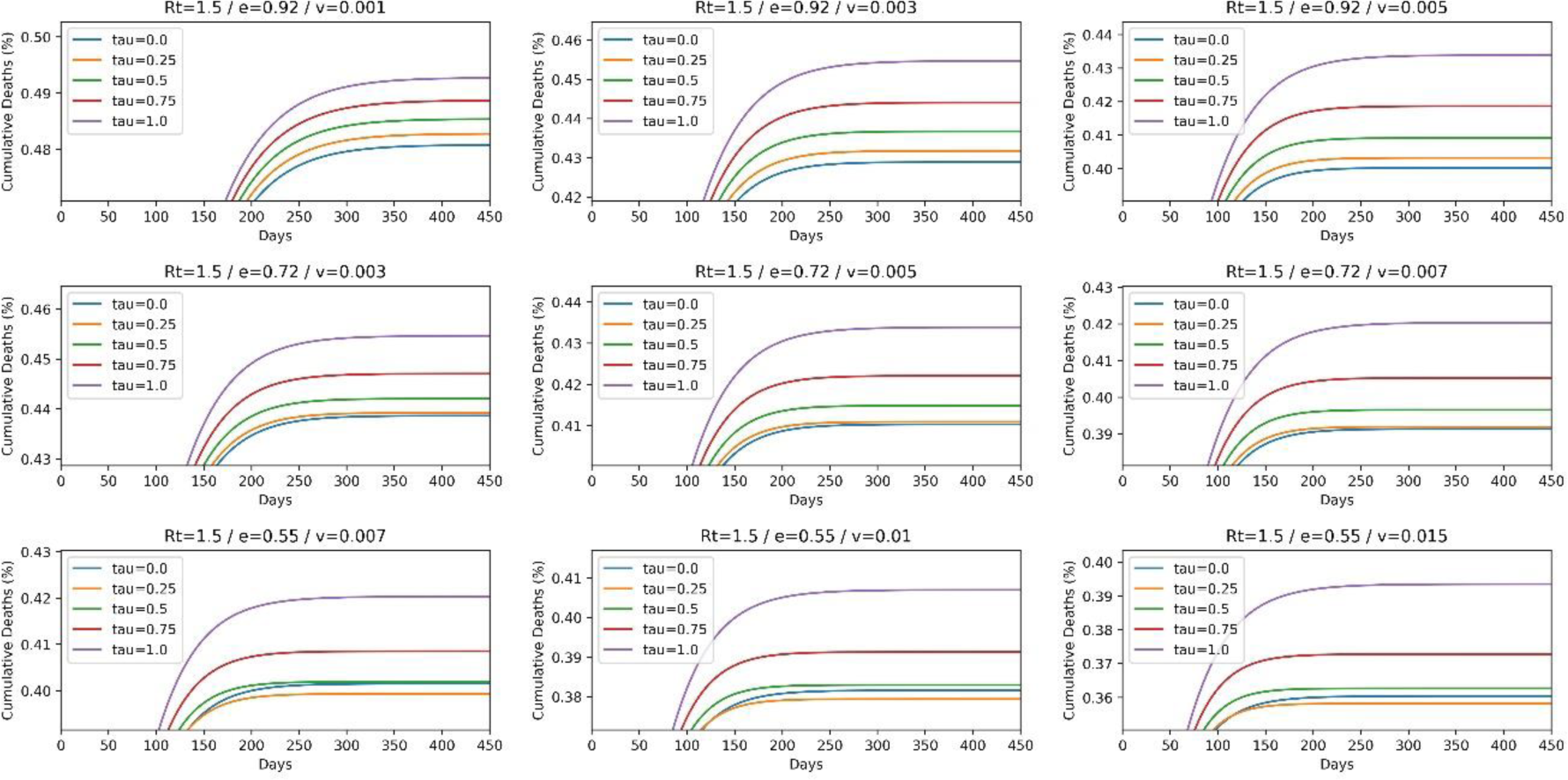
Graphs showing the percentage of cumulative deaths in a population with infection characteristics similar to those of Russia with a transmission coefficient of R=1.5 with initial conditions: 1.84% currently infected, 27.12% currently immune, 22.62% currently susceptible with lost immunity, 0.23% currently dead. Data collected using the following 9 vaccination schemes: e=92% and v=0.1%, e=92% and v=0.3%, e=92% and v=0.5%, e=72% and v=0.3%, e=72% and v=0.5%, e=72% and v=0.7%, e=55% and v=0.7%, e=55% and v=1%, e=55% and v=1.5%, with 𝑣_𝑖_ = 90%.

In addition to that, the figure also shows that the roll out rate of the vaccine has an important impact on the percentage of cumulative deaths. For any given efficacy of the vaccine, when the rollout rate increases, the cumulative deaths decrease.

To accurately study the dynamics of the spread of COVID-19 in a population, the percentage of active cases should be measured. By using the same initials conditions used in Fig. (2), the percentage of active cases can be predicted in populations with high infection and vaccination rates like Russia.

According to Fig. (3), all vaccination schemes reach a peak in active cases shortly after the start of the simulation, then gradually decrease to reach approximately 0 after nearly 200 days. In order to minimize the number of active cases, the optimal value for tau needed is 𝜏 = 0.5. This is interesting since 𝜏 = 0.5 does not correspond to the least percentage of cumulative deaths in Fig (2).

**Figure 3.**
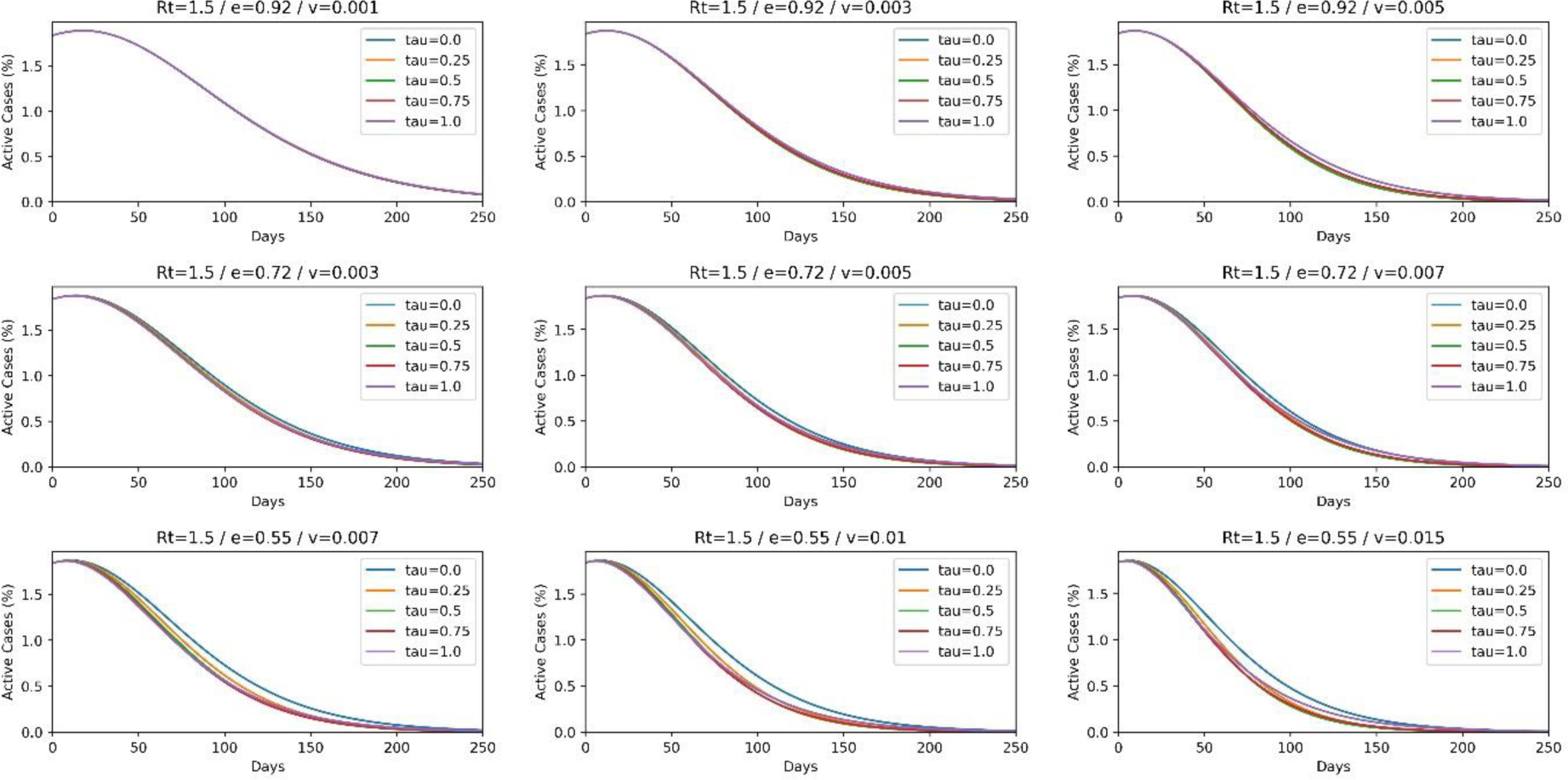
Graph showing the percentage of active cases in a population similar to Russia. Initial conditions and vaccination schemes are the same as those used in Fig. (2).

**Figure 4.**
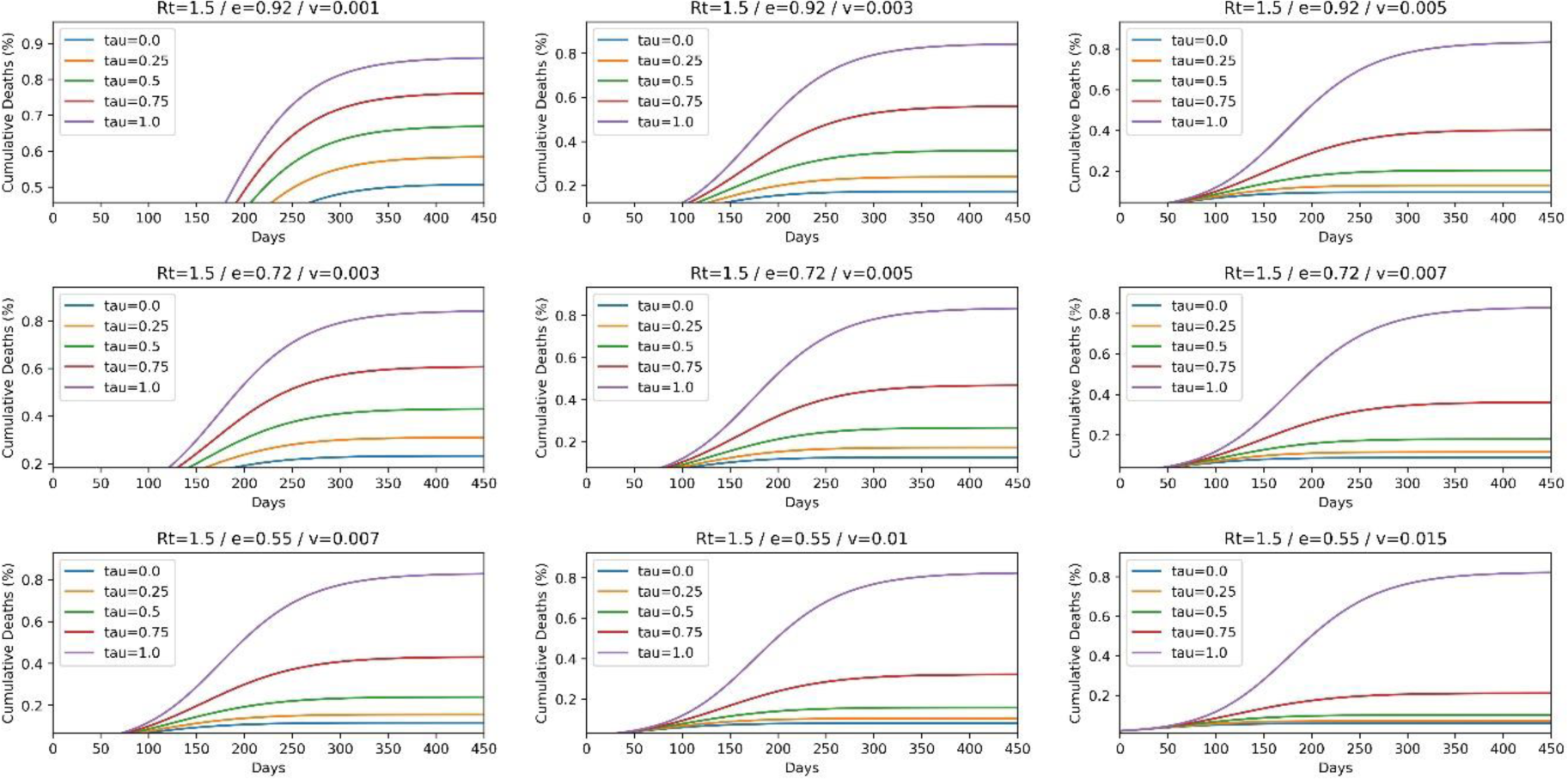
Graph showing the percentage of cumulative deaths in a population similar to that of Djibouti with a transmission coefficient of R=1.5 with initial conditions: 0.18% currently infected, 7.64% currently immune, 2.33% currently susceptible with lost immunity, 0.02% currently dead. The data collected was collected using the same vaccination schemes as Fig. (2).

In Djibouti, there was a total of 2,017 active cases in early April 2022. Therefore, with a total population of 1.1 million, the percentage of currently infected individuals is 0.18%. At that time, COVID-19 was the cause of 189 deaths in Djibouti, or 0.02% of the population. In April, 9.97% of the population was fully vaccinated, but 6 months prior, there were 2.33% double vaccinated individuals; therefore 2.33% of the population have probably lost their immunity after vaccination, leaving 7.64% that are currently immune [30, 32].

In contrast to the graphs obtained in Fig. (2), all the graphs in Fig. (3) show the same trend, which is that when the proportion of vaccine doses used for third doses decreases, the percentage of cumulative deaths also decreases. Consequently, the optimal value of tau which achieves the lowest percentage of deaths is 𝜏 = 0; However, Fig. (2) and (3) share one finding, which is that as the roll out rate of vaccines increases, the percentage of death decreases.

By graphing the percentage of active cases in a population similar to Djibouti,the following graphs are obtained in Fig. (5).

**Figure 5.**
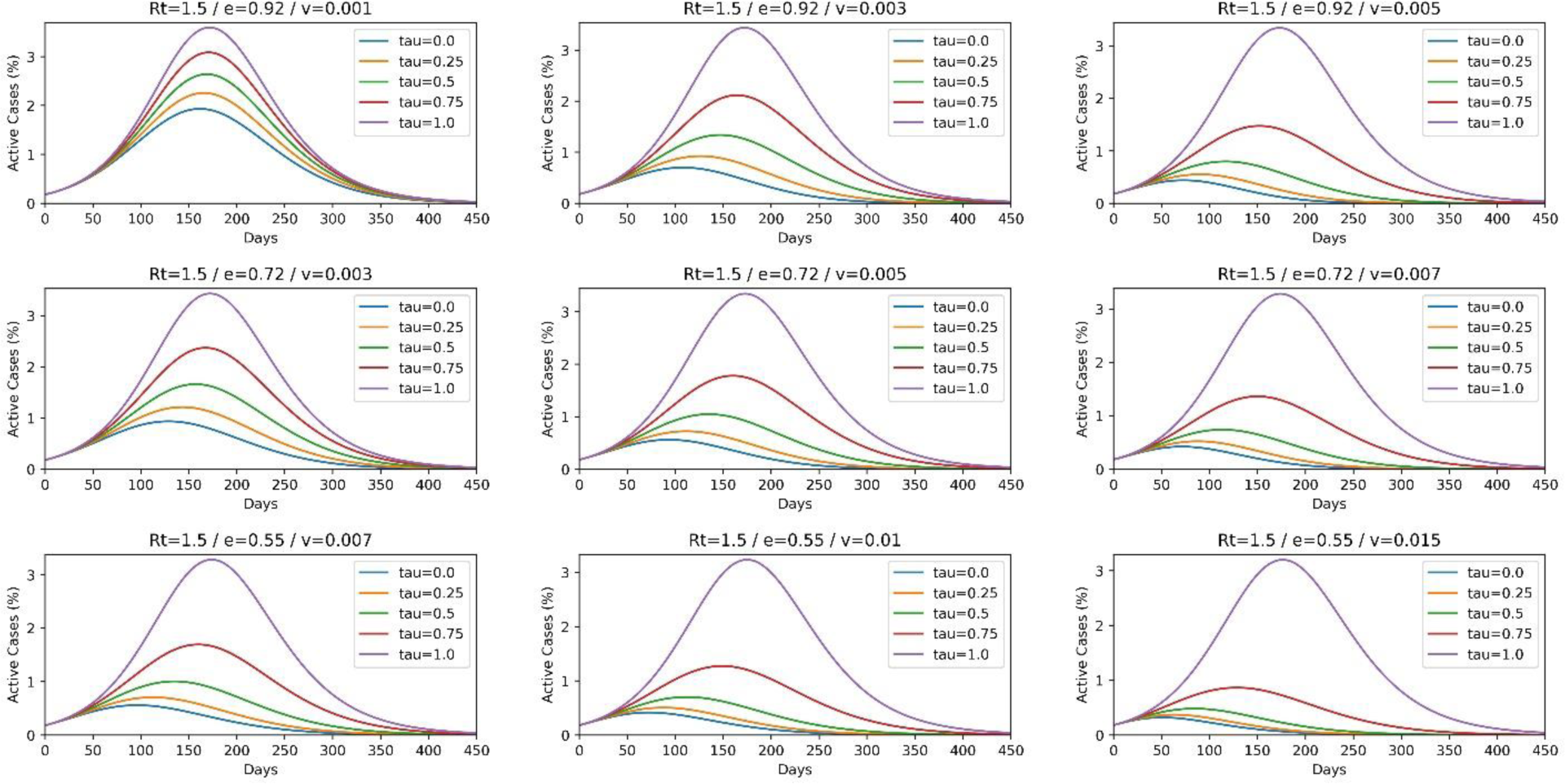
Graph showing the percentage of active cases in a population similar to that of Djibouti. The data collected was collected using the same initial conditions and vaccination schemes as Fig. (4).

Fig. (5) shows that for 𝜏 = 1, in all vaccination schemes, the percentage of active cases reaches a peak around 200 days reaching around 3% of the total population. However, as the proportion of vaccines used for third doses decreases, the peak of the percentage of active cases decreases in amplitude and is shifted to the left. Therefore, it can be concluded that for a population with a low infection and vaccination rate, the optimal way to reduce active cases is by using all the available doses to give two doses to unvaccinated individuals.

In addition to that, roll out rate seemed to play an important role in decreasing the peak of active cases. As seen in the bottom-right graph which corresponds to a vaccination scheme with a 55% efficacy and the highest roll out rate of 1.5%, the peak, for the optimal tau value of 𝑟 = 0, the peak was obtained at around day 50 with an amplitude of approximately 0.3%.

These findings are crucial in order to minimize the burden of infectious diseases like COVID-19 on medical care sectors. As mentioned previously, around 2% of COVID-19 patients are hospitalized [7], so by decreasing the amplitude of the peaks, there will be less hospitalized patients at any given moment.

## 5. Limitations

In this study, we introduced a novel model to simulate the expected results of using vaccine shots as boosters or as first doses. However, in this *Booster* model, there are still gaps and limitations that stand in the way of this model accurately representing the dynamics of infectious spreads in certain communities.

The maximum possible vaccine intake percentage is an important factor in analyzing and forecasting infection and death scenarios related to COVID19 and its related vaccination programs. In this study, we assumed that we are dealing with populations that would achieve around 90% vaccination rate. Though this was achieved across many countries across the world, it would be important to further continue this study and simulate different values for this parameter. This would account for populations that may be affected by anti-vaccination rhetoric and campaigns, or for poorer countries that may not have the logistics to widely spread the vaccine. A separate simulation for 𝑣_𝑖_ could be the goal of future studies.

Another important factor is that the transmission coefficient is also sensitive on mitigation measures and may vary depending on specific implementations of social distancing, cleanliness, face masks and other mitigation schemes.

Finally, tackling specific vulnerable populations like the elderly of those carrying chronic diseases specifically with booster shots in case of limited resources could also prove effective enough in minimizing deaths. This alternative was not addressed in our study.

## 6. Conclusions

The new *Booster* model allowed us to study the effect of the third dose on a community. It allowed us to find the optimal percentage of COVID-19 doses that should be administered as booster shots, rather than being used as first or second shots for unvaccinated individuals.

The obtained results show that for communities with a relatively high number of individuals that have lost their immunity after vaccination or after recovery from infection, allocating around 25% of the vaccines as third doses should lead to the lowest percentage of cumulative deaths. On the other hand, a population with a relatively low portion of individuals with lost immunity would find more benefit in vaccinating the susceptible unvaccinated individuals, rather than reimmunizing those who have done the first and second doses.

Our model and results have important potential for tackling real world problems and informing policy makers on efficient strategies. In our prototype example, we could advise health officials in one country to dedicate a quarter of their vaccine resources for boosters in case there is still need for first doses while calling for full dedication to first doses in another country with different infection conditions. In this sense, this model is generic and flexible and could be implemented to reach optimal distribution scenarios under different needs.

## Data availability statement

All the data used in the article is publicly available and referenced in the article.

## Author Contributions

EE J. performed the simulations and coding and wrote the first draft. ED O. designed the study and wrote the final draft.

## Data Availability

All data produced in the present study are available upon reasonable request to the authors

## References

1. Worldometer. (2023, July 31). COVID-19 CORONAVIRUS PANDEMIC. Retrieved July 31, 2023, from https://www.worldometers.info/coronavirus/

2. CDC. (2021, December 1). SARS-CoV-2 Variant Classifications and Definitions. Retrieved from Centers for Disease Control and Prevention: https://www.cdc.gov/coronavirus/2019-ncov/variants/variant-classifications.html?CDC_AA_refVal= https://www.cdc.gov%2Fcoronavirus%2F2019-ncov%2Fvariants%2Fvariant-info.html

3. [3] Pulliam, J. R. C., Van Schalkwyk, C., Govender, N., Von Gottberg, A., Cohen, C., Groome, M., Dushoff, J., Mlisana, K., & Moultrie, H. (2022). Increased risk of SARS-CoV-2 reinfection associated with emergence of Omicron in South Africa. Science, 376(6593). 10.1126/science.abn4947

4. British Society for Immunology. (2021, January). A guide to vaccinations for COVID-19. Retrieved from British Society for Immunology: https://www.immunology.org/sites/default/files/BSI_GuidetoVaccinationsCOVID19_Aug21.pdf

[5] Heida, R., hinrichs, W. L., & Frijlink, H. W. (2021, August 25). Inhaled vaccine delivery in the combat against, Expert Review of Vaccines. 21(7), 957–974. 10.1080/14760584.2021.1903878

6. The Economist. (2021, June 19). A new weapon in the war against SARS-CoV-2 has been found. Retrieved from The Economist: https://www.economist.com/science-and-technology/2021/06/16/a-new-weapon-in-the-war-against-sars-cov-2-has-been-found

[7] Self, W. H., Tenforde, M. W., & Arter, O. G. (2021, September 24). Comparative Effectiveness of Moderna, Pfizer-BioNTeck, and Janssen (Johnson & Johnson) Vaccines in Preventing COVID-19 Hospitalization Among Adults Without Immunocompromising Conditions - United States, March-August 2021. Retrieved from Morbidity and Mortality Weekly Report: 10.15585/mmwr.mm7038e1

[8] Menachemi, N., Dixon, B. E., Wools-Kaloustian, K., Yiannoutsos, C. T., & Halverson, P. K. (2021). How Many SARS-CoV-2–Infected People Require Hospitalization? Using Random Sample Testing to Better Inform Preparedness Efforts. Journal of Public Health Management and Practice, 27(3), 246–250. 10.1097/phh.0000000000001331

[9] Kermack, W., & McKendrick, A. (1927). A contribution to the mathematical theory of epidemics, Proceedings of the Royal Society A, 115(772).

[10] Melis, M., & Littera, R. (2021). Undetected infectives in the Covid-19 pandemic. International Journal of Infectious Diseases, 104, 262–268.

[11] Negrut, N., & Ivan, T. (2021), Long COVID-19 – a pathology of concern. International Journal of Infectious Diseases, 116, 547. 10.1016/j.ijid.2021.12.113.

[12] El Deeb, O., Hattaf, K., & Kharroubi, S. (2023), Modeling of COVID-19 and other infectious diseases: Mathematical, statistical and biophysical analysis of spread patterns, Frontiers in Applied Mathematics and Statistics, 9:1178479. 10.3389/fams.2023.1178479.

[13] Hattaf, K., Mohsen, A. A., Harraq, J., & Achtaich, N. (2021). Modeling the dynamics of COVID-19 with carrier effect and environmental contamination. Advances in Complex Systems, 12(03), 2150048. 10.1142/s1793962321500483.

[14] Giandhari, J., Pillay, S., Wilkinson, E., et al., Early transmission of SARS-CoV-2 in South Africa: An epidemiological and phylogenetic report (2020), International Journal of Infectious Diseases, 103, 234–241. 10.1016/j.ijid.2020.11.128.

[15] El Deeb, O., & Jalloul, M. (2020, August 19). The dynamics of COVID-19 spread: evidence from Lebanon, Mathematical Biosciences and Engineering, 17(5), 618–5632. doi:10.3934/mbe.2020302

[16] Kharroubi, S. A., & Saleh, F. (2020). Are lockdown measures effective against COVID- 19? Frontiers in Public Health, 8, 549692. 10.3389/fpubh.2020.549692.

[17] Kim., Y.J., Seo, M.H. & Yeom, H.E., Estimating a breakpoint in the pattern of spread of COVID-19 in South Korea (2020), International Journal of Infectious Diseases, 97, 360–364. 10.1016/j.ijid.2020.06.055.

[18] Kharroubi, S. (2020), Modeling the Spread of COVID-19 in Lebanon: A Bayesian Perspective, Frontiers in Applied Mathematics and Statistics, 6:40. doi: 10.3389/fams.2020.00040.

[19] Youssef, D., Issa, O., Kanso, M., Youssef, J., Abou-Abbas, L., & Abboud, E. (2022). Practice of non-pharmaceutical interventions against COVID-19 and reduction of the risk of influenza-like illness: a cross-sectional population-based study. Journal of Pharmaceutical Policy and Practice, 15(54). 10.1186/s40545-022-00450-y

[20] Youssef, D., Berry, A., Youssef, J., & Abpu-Abbas, L. (2022), Vaccination against influenza among Lebanese health care workers in the era of coronavirus disease 2019, BMC Public Health, 22:120. 10.1186/s12889-022-12501-9.

[21] El Deeb, O., & Jalloul, M., (2020), Forecasting the outbreak of COVID-19 in Lebanon, medRxiv, 10.1101/2020.09.03.20187880.

[22] Hattaf, K. & Yousfi, N., (2020), Dynamics of SARS-CoV-2 infection model with two modes of transmission and immune response, Mathematical Sciences and Engineering, 17(5), 5326–5340. Doi:10.3934/mbe.2020288.

[23] Abou Hassan, F., Bou Hamdan, M., Ali, F., & Melhem, N. (2023), Response to COVID-19 in Lebanon: update, challenges and lessons learned, Epidemiology and Infection, 151:23. 10.1017/S0950268823000067.

[24] El Deeb, O., & Jalloul, M. (2021, September 2). Efficacy versus abundancy: Comparing vaccination schemes, PLoS ONE 17(5): e0267840. 10.1371/journal.pone.0267840

[25] Collier, A. Y., Yu, J., McMahan, K., Liu, J., Chandrashekar, A., et al., (2021). Differential Kinetics of Immune Responses Elicited by Covid-19 Vaccines. The New England Journal of Medicine, 385(21), 2010–2012. 10.1056/nejmc2115596

26. Mishra, M., & Erman, M. (2021, August 14). U.S. authorizes third shot of COVID-19 vaccines for the immunocompromised. Reuters. https://www.reuters.com/world/middle-east/us-fda-authorizes-covid-19-vaccine-boosters-immunocompromised-2021-08-13/

27. [27] The Connexion. (2021). France to give third Covid vaccine dose to those with immunodeficiency. https://www.connexionfrance.comwww.connexionfrance.com/article/French-news/France-to-give-third-Covid-vaccine-dose-to-those-with-immunodeficiency

[28] Krause, P. R., Fleming, T. R., Peto, R., Longini, I. M., Figueroa, J. P., & Sterne, J. A. (2021, October 09). Considerations in boosting COVID-19 vaccine immune responses. The Lancet, 398: 10308, 1377–1380. 10.1016/S0140-6736(21)02046-8

[29] Theparod, T., Kreabkhontho, P., & Teparos, W. (2023). Booster Dose Vaccination and Dynamics of COVID-19 Pandemic in the Fifth Wave: An Efficient and Simple Mathematical Model for Disease Progression. Vaccines, 11(3), 589.

30. Mathieu, E. (2020, March 5). Coronavirus pandemic (COVID-19). Our World in Data. https://ourworldindata.org/covid-vaccinations

31. [31] Worldometer. (2021). Russia COVID - Coronavirus Statistics - Worldometer. https://www.worldometers.info/coronavirus/country/russia/

32. [32] Worldometer. (2021). Djibouti COVID - Coronavirus Statistics - Worldometer. https://www.worldometers.info/coronavirus/country/djibouti/

